# Analyzing county-wide trends in Tennessee Covid-19 rates, Median Household Income, and Presence of Hospital

**DOI:** 10.1101/2022.03.02.22271656

**Authors:** Viraj Brahmbhatt

## Abstract

The Covid-19 pandemic has caused millions of deaths and infections worldwide. Recent studies suggest that Covid-19 may be disproportionately affecting certain groups. The Tennessee Department of Health regularly publishes data on Covid infections and vaccinations. This data alongside data published from the 2010 census was used to analyze trends in Covid-19 rates for the State of Tennessee. The census data for the average household income of each county was cross-referenced with the covid data. A positive correlation between population of the county and the number of new cases reported on January 3^rd^, 2022, appeared when observing the data. A regression analysis (ANOVA) revealed that the data on population and covid rate was significant (P-value: 2.69E-10). The results from comparing the covid rates in a county with a hospital and a county without a hospital seemed to be the most significant. The data reported by the Sycamore institute on the Tennessee counties without a hospital was used to identify trends unique to these counties. The counties lacking a hospital were compared with counties with a similar population. 7 hospital-less counties were used for comparison, 6 counties (Fayette, Grainger, Haywood, Chester, Sequatchie, Clay) reported a greater number of cases than counties of a similar population. Of the 20 counties lacking a hospital, 16 fell within the bottom 50% of median household incomes, with 9 in the bottom 25% and 4 in the bottom 10%. Healthcare sites in rural areas may lack fundamental infrastructure. These areas may require unique interventions to address the healthcare concerns present.

## 1. Introduction

The Covid-19 pandemic (SARS-COV-2) is characterized by an upper respiratory infection. The World Health Organization reports 308,458,509 confirmed cases of COVID-19 and 5,492,595 deaths globally due to the pandemic as of January 11^th^, 2022 [1]. A major development in the trajectory of the disease came on December 11^th^, 2020 as with the release of the Pfizer/BioNtech Covid vaccine for individuals above 16 [2]. Since this initial release, Moderna and Johnson&Johnson all released their own versions of the Covid vaccine. On August 23, 2021, the Pfizer/BioNtech Covid vaccine received FDA approval [3]. Since then, the CDC has approved the Pfizer/BioNtech vaccine have been approved to be administered to individuals above the age of 5 [4].

In certain areas of the country, there has been noticeable resistance to Covid vaccination. A 2021 publication from Edwards et al. found that those living in highly religious areas – as well as Females, those with populist views, and those who felt that Covid’s effects were not as extreme as stated – were more likely to resist Covid vaccination [5]. The term bible belt is used to colloquially refer to the region containing the 10 states, Mississippi, Alabama, Louisiana, Arkansas, South Carolina, Tennessee, North Carolina, Georgia and Oklahoma. This term came about due to conservative social attitudes present alongside a high position of religion in culture [6]. As suggested by the adherence trends discussed in the 2021 Edwards publication, this area likely displays low adherence to Covid guidelines [5].

Since the release of the Covid vaccines, the mortality rate of the disease has decreased. [7]With new variants of the SARS-COV-19 being mutated, there have been recurrent waves. The most recent wave of the Omicron variant of SARS-COV-19 has seen record numbers, surpassing all past highs of Covid – 19 cases in the state of Tennessee [8]. This rise in Covid rates prompts investigation on how different groups have been affected by Covid. Studies such as the 2021 Magesh et al. study have investigated the socio-economic factors associated with Covid outcomes. The Magesh et al. study found strong correlations between race and Covid-19 outcomes [9]. In addition to race, the financial standing of a county may also prove to be a noteworthy contributor to Covid-19 outcomes. The 2021 Tan et al. publication found a positive correlation between income inequality and Covid 19 cases and deaths for the cohort of all counties in the United States [10]. For the State of Tennessee, an investigation on the economic status (as measured through median household income provided through recent Census data) and Covid outcomes may prove to be insightful.

## 2. Methods

Data on the rates of the Covid pandemic in the State of Tennessee were obtained from the downloadable dataset portion of the Tennessee Department of health website. Microsoft Excel was used to categorize and analyze the data. The data was sorted by the date of collection. A linear regression model was used to analyze the data. January 3^rd^, 2022 was selected as the date to be studied as it was the most recent complete collection date. The data was categorized by county and sorted based on county population. The most pressing data came from comparing the counties with the greatest population and greatest active covid rates. The lack of the presence of a hospital within the county border was cross referenced with the counties with the counties with the most covid infections. The median household income obtained from 2010 Census Data was also compared to each county to investigate the correlation of county wealth and fate during recent Covid waves.

### 2.1 Counties without hospitals

To prepare the data for comparison, the counties with no hospitals were found through the sycamore institute [11]. The report indicated that there were 20 counties in the state of Tennessee without an open hospital. The names of the counties were cross referenced with the top 10 counties with the highest covid cases as well as the counties general position within the distribution of all counties by wealth.

### 2.2 Economics of Counties and Covid rates

The economic standings of the counties were determined through accessing 2010 census data. The Median Household income per county was used as an indication of the economic standing of the county. To simplify the analysis of the correlation, the top 15 counties in each category were used for trend analysis. Specifically, the counties in the greatest 15 and lowest 15 Median Household Income were cross referenced. Additionally, counties lacking a hospital were cross referenced with their corresponding Median household income.

### 2.3 Vaccination Percentage and Median Household Income

Vaccination percentage, a measurement of the ratio of individuals within the county that are vaccinated, was obtained through taking the number of reported vaccinated individuals and dividing this figure by the total population of the county. It is important to note that this figure is not completely accurate. The population data and the vaccination numbers were obtained from separate data banks from the Tennessee Department of Health online data. The inaccuracies in the data were especially present with certain counties in which the vaccination percentage was greater than 100%. These counties were removed from the comparison as it is likely that reported data from these counties was not accurate. Further analysis investigated vaccination percentage that were not likely but were less than 100%. It is reported that ∼55% of Tennessee’s population is fully vaccinated. As such, counties with a vaccination percentage greater than 55% were removed from the comparison as they either possessed inaccurate vaccinations rates or they were anomalous in comparison to the general state population. As such, these counties were deemed to be capable of skewing the data and were removed. The remaining data contained 75 counties, with 20 having been removed due to data that was deemed extraneous.

## 3. Results

A positive correlation between population of the county and the number of new cases reported on January 3^rd^, 2022 appeared when observing the data. The data presented a strong positive linear relationship (R^2^=.9173) between a county’s population and new covid cases documented. The two variables also showed strong correlation in regression analysis (ANOVA). The p value reported was 3.98E-52. Given that the p-value is less than .05, the relationship of correlation observed is not likely due to chance.

A similar comparison may be derived in looking into the population of a county and the number of fully vaccinated individuals (presented in Figure 2.). This data presents a weaker correlation than seen in Figure 1. It is important to note that only individuals that were fully vaccinated as of January 3^rd^, 2022 were included in the data. This data did not consider whether these individuals had received a booster dose. Additionally, partially vaccinated individuals were also not included in the data presented. The data in Figure 2 seems to imply that there is a weakly linear relationship between population of a county and the number of Fully vaccinated individuals. A regression analysis (ANOVA) was conducted on the data. The p-value for this data set was 2.69E-10. Given that the p-value is less than .05, the relationship of correlation observed is not likely due to chance and the null hypothesis is rejected.

**Figure 1.**
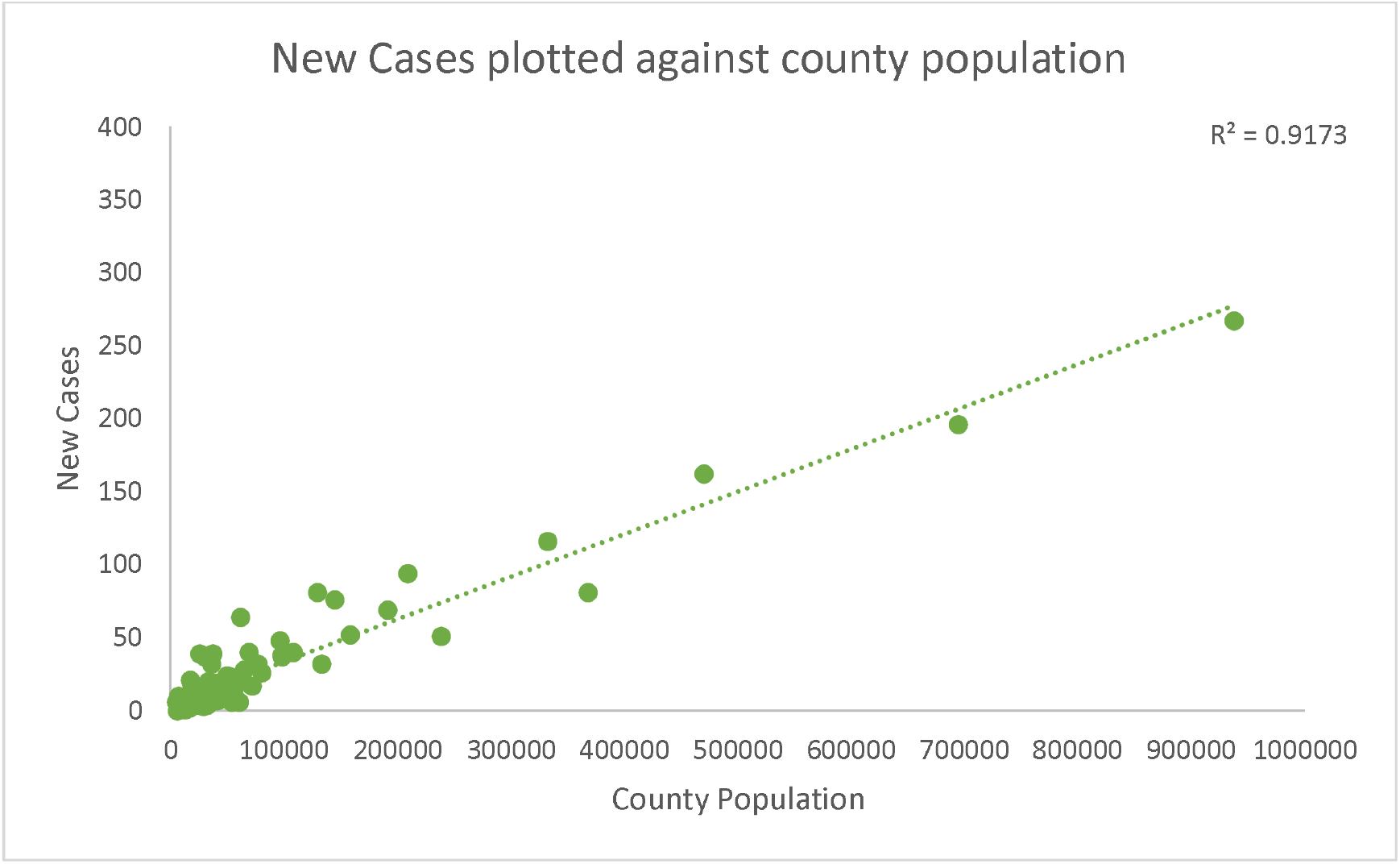
New Covid Cases plotted against population of county.

**Figure 2.**
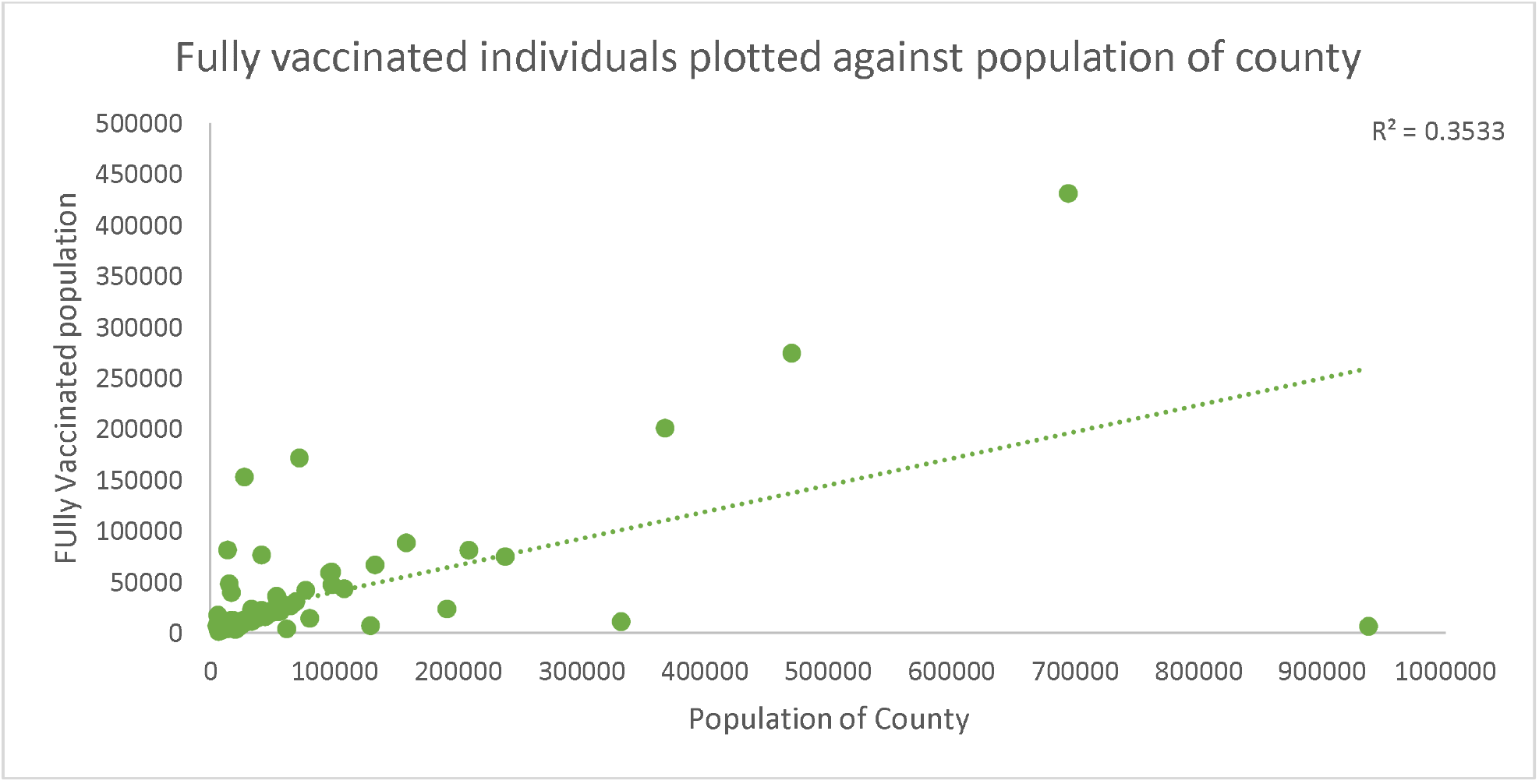
Number of fully vaccinated individuals plotted against the population of the counties.

### 3.1 Counties Without Hospitals

The data reported by the Sycamore institute on the Tennessee counties without a hospital was used to identify trends unique to these counties. The counties which lacked a hospital were compared with counties with a similar population. The Sycamore institute reported that there were 20 counties that lacked a hospital. The counties were sorted by population in each, if a successive county had a smaller population but a greater number of covid cases, then the county was flagged. The list of flagged counties was cross referenced with the counties without a hospital. 11 of the counties without a hospital were present on the flagged list. Of these 11, 7 were noted as being relevant for comparison (Fayette, Grainger, Haywood, Chester, Sequatchie, and Clay). Counties without a hospital generally reported a greater number of new covid cases compared to counties with a similar population size. Of the 7 counties available for comparison, 6 counties (Fayette, Grainger, Haywood, Chester, Sequatchie, Clay) reported a greater number of cases.

**Table 1.** County comparisons are shown. The second county in each comparison is the county lacking a hospital

| County | Population | New Covid Cases |
| --- | --- | --- |
| Warren | 41283 | 7 |
| Fayette | 41129 | 8 |
| Macon | 24597 | 7 |
| Grainger | 23315 | 9 |
| Johnson | 17784 | 5 |
| Haywood | 17327 | 10 |
| Johnson | 17784 | 5 |
| Chester | 17304 | 21 |
| Bledsoe | 15062 | 3 |
| Sequatchie | 15033 | 4 |
| Perry | 8078 | 2 |
| Clay | 7622 | 4 |
| Perry | 8078 | 2 |
| Lake | 7011 | 10 |

Using Census data, the median household income was obtained and cross referenced with the list of counties lacking a hospital. The counties were sorted by median household income, a measure used to estimate economic standing. This list was then cross referenced with the counties flagged for lacking a hospital and possessing disproportionate numbers of covid cases relative to population. The notable trends were derived through analyzing where in the median household income distribution the hospital less counties fell. Of the 20 counties lacking a hospital, 16 fell within the bottom 50% of median household incomes, with 9 in the bottom 25% and 4 in the bottom 10%.

### 3.2 Vaccination Percentage and Median Household Income

. A linear regression as well as correlation analysis was conducted. The data was weakly linear (R^2^ = .046) and did not present a strong correlation (p-value = .064). These values suggest that the correlation between Average Median Household income and covid vaccination percentage are only weakly correlated. The high p-value suggests that there may be an alternate explanation for the two variables.

## 4. Conclusions and Discussions

The most significant data analyzed came from comparing counties with similar sized populations (but with one lacking a hospital). The data suggested that while comparing two counties of similar population sizes, the county lacking a hospital was likely to have more covid cases. The implication here is that presence of a hospital within the county is a major indicator of the health of the citizens living within the county. A 2020 publication from Hawkins, Charles, and Mehaffey discovered the link between socioeconomic status and Covid-19 related fatalities. The findings from this study revolved around race and education. The Anderson et al. publication from the BMC Heath Services Research found that rural residents had lower quality health than their urban counterparts [12]. The results of studies such as these propose a duality in health quality. For the State of Tennessee, 93% of the population is living in a rural area [13]. The Covid-19 pandemic has brought about a new set of challenges to providing quality of care. In the state of Tennessee, positive covid testing rates persist, despite the push to provide vaccination. Tennessee has 11% fully vaccinated individuals than the national average. In the past 14 days (1/15/22 – 1/29/22), the national hospitalization rate fell 2%, while the Tennessee hospitalization rate due to covid was up 19% [14]. The results of this study found that the negative effects of Covid (measured through number of new infections and number of vaccinations) were more severe in counties that lacked a hospital. Additionally, the number of new cases had a strong correlation with county population (R^2^ = .9173). The number of fully vaccinated individuals, however, was only weakly correlated with county population (R^2^ = .3533). A further analysis of the counties without a hospital revealed that many of these counties ranked in the bottom 25% of counties by median household income. Additionally, when counties lacking a hospital were compared with counties with similarly sized populations, the counties lacking a hospital typically showed more severe negative effects of COVID-19 (as measured through new cases). This seems to be the most notable finding of the study. The presence of a hospital within a community is likely to improve community health through easy access; however, it may also change the health outcomes in the community. The cause behind this community health improvement may be the topic of further research. Certain studies have investigated political identification within a county and response to Covid-19 guidelines [15]. Tennessee is regarded as a politically red state or that it has voted republican in the past 3 presidential elections (with notable exceptions in the democratic counties of Davidson, Haywood, and Shelby). This factor of social identity may play a role in shaping the response to covid guidelines. As such, this factor may present itself as confound to the implication presented through comparing the data between a county without a hospital with a county lacking a hospital.

## 5. Implications and Closing Statement

The correlation seen in the data between covid rates and lack of the presence of a hospital presents a case for intervention by authorities. In the modern healthcare scene, rural areas may lack the infrastructure needed to set up proper healthcare sites. These rural residents may need healthcare interventions more than their urban counterparts. The 21^st^ century has seen healthcare grow beyond large hospitals. Ingenuity will be required to address the concerns of adequate rural healthcare. Remote clinics, growing the role of the Nurse Practitioner, or encouraging the rise of government clinics in these underserved areas may prove to be effective.

### 5.1 Vaccination Percentage and Median Household Income

The weak correlation between the vaccination percentage and Average Median household income suggested the presence of other factors. The general sentiment towards Covid protocol and guidelines may be an important factor influencing the vaccination rates. Given the political composition of Tennessee and the results from previous studies suggesting the influence of such factors on Covid protocol adherence, the vaccination percentages may be significantly influenced by these factors. The effect would likely be that the correlation to median household income would present weaker as there are more factors influencing the data.

A major limitation of the study is the reliability of the data. The data from the Tennessee department of health was recent and is vulnerable to having errors attributable to rushed data collection. The Census Data came from 2010, which is older than the other data collected. However, the data used from the census was the median household income. This figure is not likely to have changed significantly from the 2010 levels. Additionally, changes in the incomes were likely generally proportional for all the population (inflation). The general trend analysis conducted using the median household income looked at comparative levels among counties for analysis.

### 5.2 Future Research

Future Studies may choose to investigate the healthcare systems that cover the areas that have been affected by covid the greatest. The methodology used in this study may be used for a different state. Additionally, the results of this study may be cross-referenced with the racial composition of the counties that lack a hospital. The methods applied in this study may also be extrapolated to other points during the timeline of the virus. It may be interesting to see whether these trends were also present during the first and second major outbreaks for Covid-19. Furthermore, it may be interesting to see whether the trends of healthcare disparities hold true for other states containing a different percentage of rural residents. It may be possible that the trends become less apparent with a greater urban population.

## Data Availability

Median Household Income - 2010 Census Data
County Population and Covid-19 rates - TN Department of Health
Counties without a hospital - Sycamore Institute

https://www.tn.gov/health/cedep/ncov/data/downloadable-datasets.html

## Author’s Biography

Viraj Brahmbhatt is an undergraduate student in the Union College, Albany Medical College combined Leadership In Medicine Program. The author was born and raised in Johnson city, Tennessee and spent considerable time volunteering and shadowing at the Mountain Home Veteran’s Affairs Medical Center as well as the Johnson City Medical Center. Brahmbhatt graduated from University School of East Tennessee State University as the Valedictorian of the class of 2021. Brahmbhatt is interested in a career in surgical medicine.

